# Assessing and managing bone-health and fracture risk in Parkinson’s disease: the BONE PARK 2 protocol

**DOI:** 10.1101/2024.11.08.24316887

**Authors:** Katie C Naylor, Emma Tenison, Sarah A Hardcastle, Veronica Lyell, Celia L Gregson, Emily J Henderson

## Abstract

**Background:** In Parkinson’s disease (PD) the propensity to fall and the higher risk of osteoporosis converge yielding a high fracture risk. Updated National Osteoporosis Guideline Group (NOGG) guidance recommends that PD should trigger a risk assessment, for example using the FRAX tool, yet clinical pathways remain sub-optimal. To address this, we generated an algorithm for the assessment and management of bone-health specifically in PD.

**Methods:** Within the Proactive and Integrated Management and Empowerment in Parkinson’s Disease randomised controlled trial (PRIME-UK RCT), bone-health metrics were collected, and all participants were offered a DXA scan. The FRAX tool was used to obtain the 10-year probability of hip and major osteoporotic fracture (MOF), and the resulting NOGG risk-category recorded. Probabilities were recalculated including femoral-neck bone mineral density (FN-BMD) and/or with numeric adjustment for recurrent falls, and results compared.

**Results:** Among 182 people with parkinsonism (mean age 73.8 years, 65% male, median disease duration 5-years), 28.0% reported a prior fragility fracture, and 40.7% recurrent falls over the previous year. 28.6% had MOF above NOGG intervention thresholds (IT); whilst 12.1% had a FN-BMD T-Score ≤-2.5. Recalculation of FRAX with FN-BMD (n=182) reduced fracture MOF and hip fracture probabilities; 12 (6.6%) deescalated below the IT, and 16 (8.8%) moved above the IT.

**Conclusions:** This 2024 BONE-PARK algorithm is informed by both the latest NOGG Guidelines and novel findings in a ‘real-world’ population. The algorithm will aid bone health assessment people with PD.

**Key points:**

- Bone-health in Parkinson’s is under-recognised and under-treated, and we have demonstrated an ongoing treatment gap.
- In this population, we have demonstrated DXA is feasible but infrequently changes treatment.
- Our Parkinson’s specific guidance supports clinicians and patients to recognise, investigate and treat fracture risk.

## Introduction

The propensity to fall and increased osteoporosis risk converge yielding a high risk of fractures in people with Parkinson’s Disease (PD). Osteoporotic fracture risk (particularly at the hip) is more than doubled in PD and, in women of a given age, PD has been reported as the strongest single contributor to fracture risk [1,2]. After a fracture, people with PD are more likely than controls to develop complications [3,4], have difficulty regaining mobility [5] and mortality is doubled [6]. Hip fracture admissions account for 4.2% of all PD admissions in England, with an average yearly cost estimated at £13.7 million [7].

In PD neuropsychiatric symptoms, musculoskeletal and gait dysfunction contribute to high fall and fracture risk [8]. Additionally, swallowing difficulties and cognitive impairment can preclude treatment with oral osteoporosis medications. Current evidence is poorly generalisable as studies fail to capture the heterogeneity of PD, the full age spectrum nor people with cognitive impairment through restricted eligibility criteria [9,10].

Patients and clinicians are often unaware of the excess fracture risk resulting in a major treatment gap. Patients often underestimate their fracture risk [11]. Despite recent meta-analyses highlighting the elevated risk of fragility fracture in PD, particularly at the hip [12], clinical practice remains suboptimal. In 2020 Parkinson’s-UK reported that of 1131 patients assessed, 50% had fallen, yet only 1-in-6 were on bone-health treatment, and 73% lacked an up-to-date fracture risk assessment [13]. This highlights how bone-health is commonly overlooked in clinical practice.

Since 2019, when we published the ‘BONE-PARK algorithm’ [14], which updated our 2014 guidance [15], and has been used as the gold standard for the Parkinson’s-UK service improvement project [14], there have been two significant developments in the management of bone-health. In September 2021, new National Osteoporosis Guideline Group UK (NOGG) guidelines were launched, which specifically mention PD as a risk factor for fracture that should trigger risk assessment, for example using the FRAX tool [16,17]. Furthermore, NOGG provides new post-FRAX fracture risk categories, which stratify patients to guide management decisions.

FRAX is a fracture risk assessment tool that incorporates risk factors to predict the 10-year probability of sustaining a major osteoporotic fracture (MOF) or hip fracture. Bone mineral density (BMD), measured by Dual X-Ray Absorptiometry (DXA), can be included in FRAX. This is advantageous as, whilst osteoporosis is operationally defined using BMD alone (T-Score≤-2.5), most fragility fractures occur in people who do not meet this criterion [18,19]. However, a key limitation is that falls are not directly incorporated into FRAX. This is important since 60% and 39% of people with PD experience falls and recurrent falls respectively [20]. Rather, FRAX assumes an average exposure to falls in the previous year despite falls being an independent risk factor for fracture [21–23], meaning fracture risk is likely under-estimated in those with elevated falls risk.

To address these evidence gaps we undertook a cross-sectional sub-study of people with parkinsonism to determine fracture risk in a high risk, under-served, representative population. Our objectives were to: 1) identify the current treatment gaps; 2) determine how including femoral neck BMD (FN-BMD) influences FRAX probabilities and subsequent NOGG-based treatment recommendations; 3) consider the influence of recurrent falls as a clinical risk factor; 4) generate a new algorithm for the assessment and management of bone-health in PD.

## Methods

### Study Design and Participants

This was a cross-sectional sub-study nested within the Proactive and Integrated Management and Empowerment in Parkinson’s Disease randomised controlled trial (PRIME-UK RCT), which is evaluating a new model of care [24]. This trial enrolled 214 people with idiopathic PD and other forms of parkinsonism recruited from the catchment area of the Royal United Hospitals Bath NHS Foundation Trust (RUH Bath). Ethical approval was granted by London-Harrow Research Ethics Committee (REC reference 21/LO/0387) on 14/7/2021. Further details of the trial methodology have been previously published [24].

### Assessment of bone health

Bone-health metrics, including FRAX risk factors, were collected as part of the baseline visit [16], along with the Clinical Frailty Scale (CFS) and Movement Disorder Society-Unified Parkinson’s Disease Rating Scale (MDS-UPDRS), a measure of PD severity. Height (nearest mm, SECA stadiometer) and weight (nearest 0.1kg, SECA scales) were measured. If measurements were not possible (e.g. severe camptocormia, inability to stand etc.), most recent height and weight were taken from routine clinical records.

All participants were offered a DXA. Our protocol included bilateral hip and lumbar spine scans (PRIME-DXA). Forearm scans were also obtained if deemed necessary by the DXA operator (due to e.g. bilateral hip replacement). If participants had a DXA as part of their standard medical care less than 2-years prior to baseline, this was included in the analysis in the absence of a PRIME-DXA. All scans were performed by a trained DXA operator, using a Hologic Discovery A (S/N85953) Densitometer (Software version: 13.5.3) at RUH Bath. Daily quality control scans with the manufacturer-provided spine phantom, and weekly tests of reproducibility and tabletop radiographic uniformity were performed.

T-scores compare measured BMD to mean BMD of a young healthy reference population to diagnose osteoporosis. T-score site, calculation and interpretation followed international and national guidelines (Appendix.1) [25–27].

The 10-year probabilities of sustaining a MOF or hip fracture were calculated using FRAX (https://www.fraxplus.org/calculation-tool; accessed June 2024) [28], excluding and including FN-BMD (in g/cm2); where both hips had been scanned, the lowest of the two values was used. PD was included using rheumatoid arthritis (RA) as a proxy risk factor, as recommended by NOGG [29].

The resulting NOGG fracture risk category (‘NOGG-category’), based on FRAX probabilities, age-specific intervention and risk thresholds, was recorded. Without BMD, MOF risk is categorised as: low, intermediate, high or very-high. The intermediate category was subdivided into above or below the intervention threshold. With inclusion of BMD, MOF and hip fracture risks were categorised as: low, high or very-high [17].

### Assessing current treatment gaps

We extracted data from hospital electronic prescription records, rheumatology outpatient/ osteoporosis day unit letters and DXA reports, for documented bone protective medication prescriptions and compared the NOGG treatment recommendations. Self-reported medications were used where electronic records were not available. Participants were considered ‘on treatment’ if bone protective medication had been started and not discontinued before the baseline visit (Appendix.2).

Bone protective treatment was recommended if FRAX probabilities without BMD were greater than the NOGG intervention threshold. After inclusion of BMD, treatment was indicated if either hip fracture or MOF risk was categorised by NOGG as ‘high’ or ‘very-high’.

### Assessing the clinical value of DXA

FRAX-derived fracture probabilities and NOGG-categories, calculated excluding and including BMD, were compared. The NOGG-category for MOF excluding BMD was compared to the NOGG-category including BMD, for either MOF or hip fracture depending on which had the highest risk category [17,30].

### Assessing the influence of recurrent falls

As per NOGG guidelines, FRAX probabilities were adjusted for recurrent falls (≥2 retrospectively self-reported falls in the previous year) by increasing the probability of MOF and hip fracture by 30% [17,31].

### Statistical Analysis

Stata 18 was used for analysis. Data are described using mean/standard deviation (SD) for normally distributed variables, and if skewed, using median and inter-quartile range. Given the large sample size it was appropriate to use paired t-tests to compare the different FRAX probabilities for a given individual, according to the central limit theorem [32]. Categorical variables were described using counts with percentages and compared by chi-squared testing.

### Patient and public involvement

Patient and carer representatives were involved in the design, conduct and sharing of findings from this study.

## Results

In total, 214 people with parkinsonism were recruited into the PRIME-RCT (mean age 74.5 years SD=8.2, 65.0% male), of whom 213 had bone-health information available (Table 1).

**Table 1.** Demographic and bone-health characteristics of the PRIME-UK population.

|  | Summary |
| --- | --- |
| N | 214 |
| Age, mean (SD) | 74.5 (8.2) |
| <u>Sex, n (%)</u> |  |
| Female | 75 (35.0%) |
| Male | 139 (65.0%) |
| Weight (kg), mean (SD) | 76.1 (15.9) |
| Height (cm), mean (SD) | 168.1 (9.5) |
| <u>BMI Category, n (%)</u> |  |
| Underweight | 7 (3.3%) |
| Healthy Weight | 73 (34.1%) |
| Overweight | 81 (37.9%) |
| Obese | 51 (23.8%) |
| Severely Obese | 2 (0.9%) |
| <u>Diagnosis, n (%)</u> |  |
| Parkinsons disease | 184 (86.0%) |
| Parkinsons disease dementia | 4 (1.9%) |
| Lewy Body Dementia | 14 (6.5%) |
| Progressive Supranuclear Palsy | 4 (1.9%) |
| Multiple System Atrophy | 3 (1.4%) |
| Corticobasal degeneration | 1 (0.5%) |
| Vascular parkinsonism | 3 (1.4%) |
| Other | 1 (0.5%) |
| Years lived with PD, median [IQR] | 5.0 [3.0-8.0] |
| MDS-UPDRS Part III, mean (SD) | 42.7 (19.9) |
| <u>Clinical Frailty Score, n (%)</u> |  |
| 1 - Very fit | 5 (2.4%) |
| 2 - Well | 40 (18.9%) |
| 3 - Managing Well | 43 (20.3%) |
| 4 - Vulnerable | 44 (20.8%) |
| 5 - Mildly frail | 25 (11.8%) |
| 6 - Moderately frail | 30 (14.2%) |
| 7 - Severely frail | 22 (10.4%) |
| 8 - Very severely frail | 2 (0.9%) |
| 9 - Terminally ill | 1 (0.5%) |
| <u>Hoehn & Yahr Scale, n (%)</u> |  |
| Stage 1 | 21 (9.8%) |
| Stage 2 | 67 (31.3%) |
| Stage 2.5 | 12 (5.6%) |
| Stage 3 | 64 (29.9%) |
| Stage 4 | 33 (15.4%) |
| Stage 5 | 17 (7.9%) |
| <u>Recurrent Falls ($\geq 2$ falls over previous year), n (%)</u> |  |
| No | 118 (55.1%) |
| Yes | 88 (41.1%) |
| Unknown | 8 (3.7%) |
| **<u>Bone Protective Medications, n (%)</u>** |  |
| None | 181 (84.6%) |
| Alendronate | 21 (9.8%) |
| Risedronate | 5 (2.3%) |
| Zoledronate | 5 (2.3%) |
| Denosumab | 1 (0.5%) |
| Unknown | 1 (0.5%) |
| **<u>Bone Health Data (n=213), n (%)</u>** |  |
| Previous Fragility Fracture |  |
| No | 147 (69.0%) |
| Yes | 65 (30.5%) |
| Unsure | 1 (0.5%) |
| Parental hip fracture |  |
| No | 167 (78.4%) |
| Yes | 29 (13.6%) |
| Unsure | 17 (8.0%) |
| Current Smoking |  |
| No | 212 (99.5%) |
| Yes | 1 (0.5%) |
| Taking Oral Glucocorticoids |  |
| No | 210 (98.6%) |
| Yes | 3 (1.4%) |
| Rheumatoid Arthritis (not including PD) |  |
| No | 207 (97.2%) |
| Yes | 6 (2.8%) |
| Secondary Osteoporosis |  |
| No | 194 (91.1%) |
| Yes | 19 (8.9%) |
| Alcohol |  |
| Less than 3 units/day | 174 (81.7%) |
| 3 or more units/day | 39 (18.3%) |
| Probability of MOF (%) | 17.0 (11.5) |
| Probability of hip fracture (%) | 9.1 (9.3) |

186 (87%) had a DXA scan (175 PRIME-DXA scans and 11 available from previous routine NHS care) (Appendix.3). Those who did not have a DXA scan were older by a mean of 4.6 years (p=0.002), had higher MDS-UPDRS Part III (motor) scores by an average of 18.2 points (p<0.001), and a greater proportion were frail (82.1% vs 31.7%, p<0.001) (Appendix.4). Of those who had a DXA, 12.4% had osteoporosis (T-score ≤-2.5) and 46.8% had osteopenia (low bone mass) (T-score>-2.5 and ≤-1.0). FRAX excluding and including FN-BMD, was assessed for the 182 participants who had a DXA scan (Figure 1).

**Figure 1.**
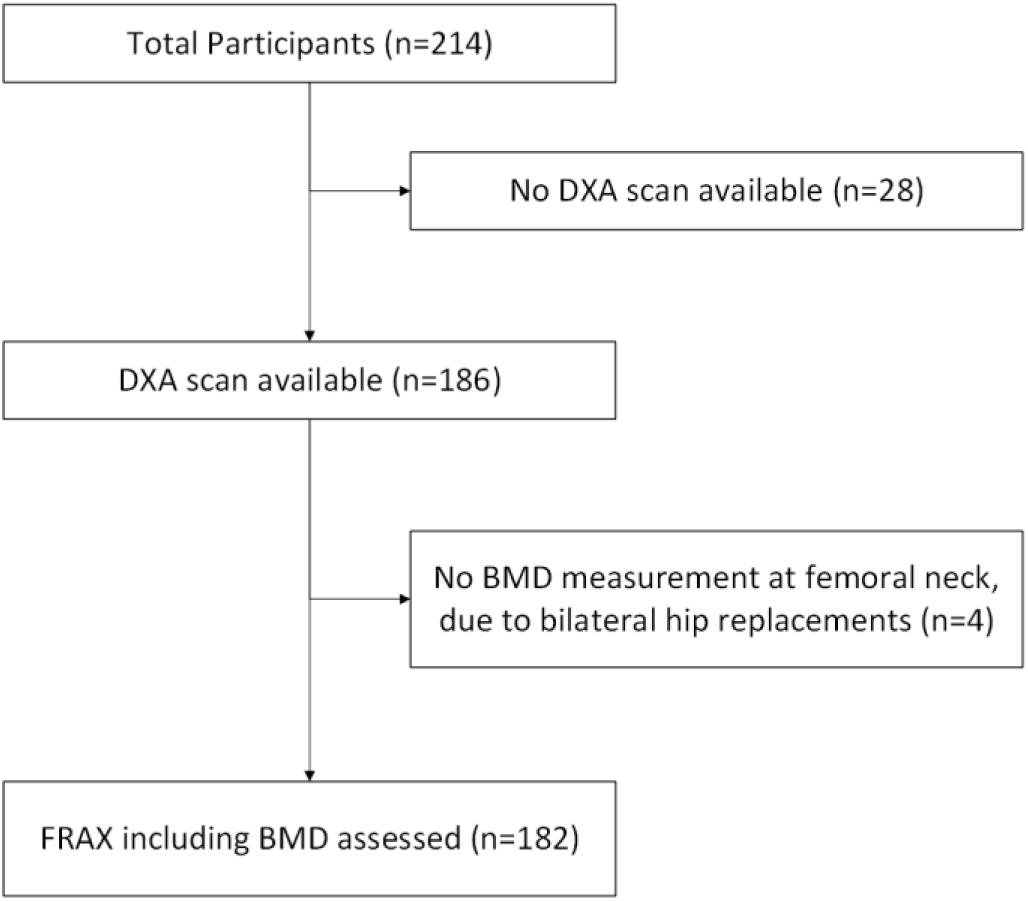
A flow chart to show how many people had FRAX assessments, with the remaining 182 forming the study population

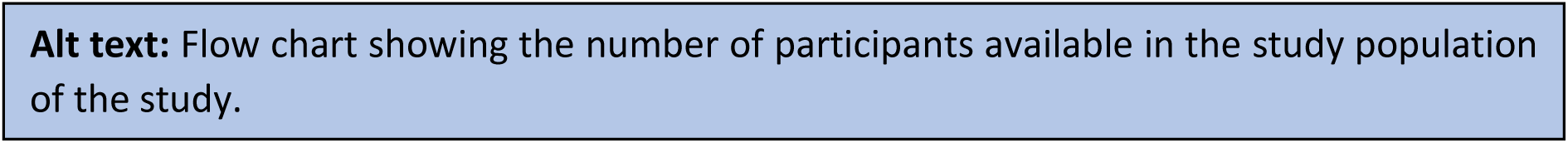

### Is there a gap between recommendation and prescribed treatment?

Only 21 participants (11.5%) were receiving bone protective treatment (Appendix.5). FRAX calculations without BMD (n=182) identified 52 (28.6%) people above the intervention threshold, of whom 15 (28.8%) were receiving bone protective treatment. FRAX recalculation including BMD (n=182) led to treatment being recommended in 56 (30.8%), of whom 15 (26.8%) were on treatment. We noted that some participants were receiving treatment despite being below the NOGG intervention thresholds (n=6 excluding BMD & n= 6 including BMD).

### What additional value does DXA add?

Recalculation of FRAX including FN-BMD reduced FRAX-derived probabilities of MOF by 3.6% (p<0.001) and hip fracture by 3.5% (p<0.001) (Appendix.6).

The effect of including BMD on the NOGG-categories is shown in Figure 2. Including BMD lowered the NOGG-category for 61 people (33.5%). This resulted in a de-escalation from above the intervention threshold (intermediate-above intervention threshold; high; or very high-risk) to lifestyle advice (low-risk) for 12 (6.6%) participants.

**Figure 2.**
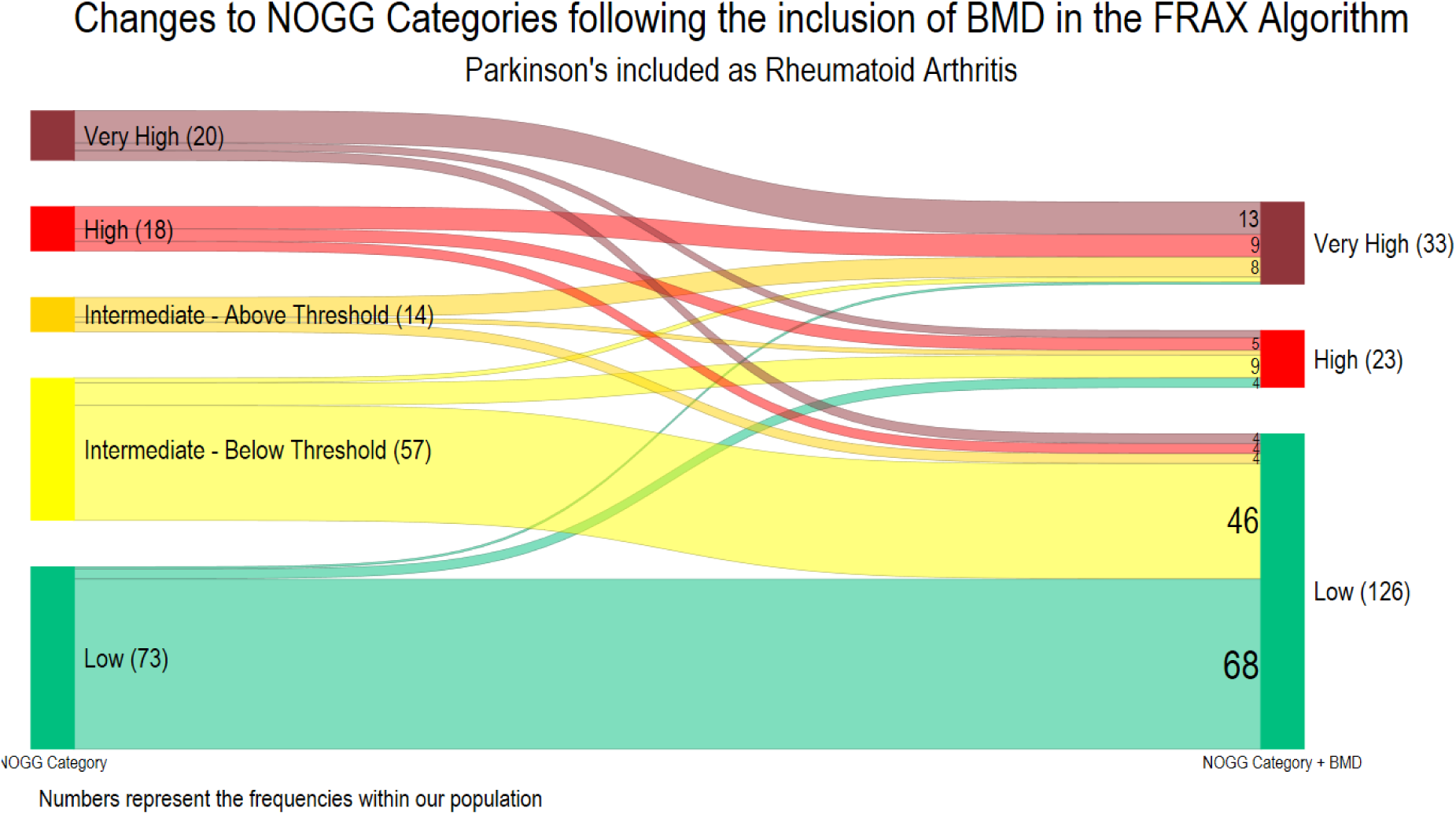
Transitions between NOGG-categories following the inclusion of femoral neck bone mineral density. A Sankey diagram to show the transition between the NOGG-categories following the inclusion of bone mineral density (BMD) into the FRAX algorithm (n=182), where PD is included as rheumatoid arthritis. Prior to the addition of BMD, the NOGG category is for major osteoporotic fracture; following the inclusion of BMD the highest NOGG category for either MOF or hip fracture was included, as the site with greatest level of risk is used to inform clinical decisions. The numbers reflect the number of participants in each category

Conversely, the NOGG-category was increased for 35 (19.2%), with an escalation from below the intervention threshold (low or intermediate-below the intervention threshold) to above the intervention threshold for 16 (8.8%) participants. Overall, the recommended management (life-style advice, treatment or treatment plus liaison with bone health specialist) remained unchanged for 134 (73.6%).

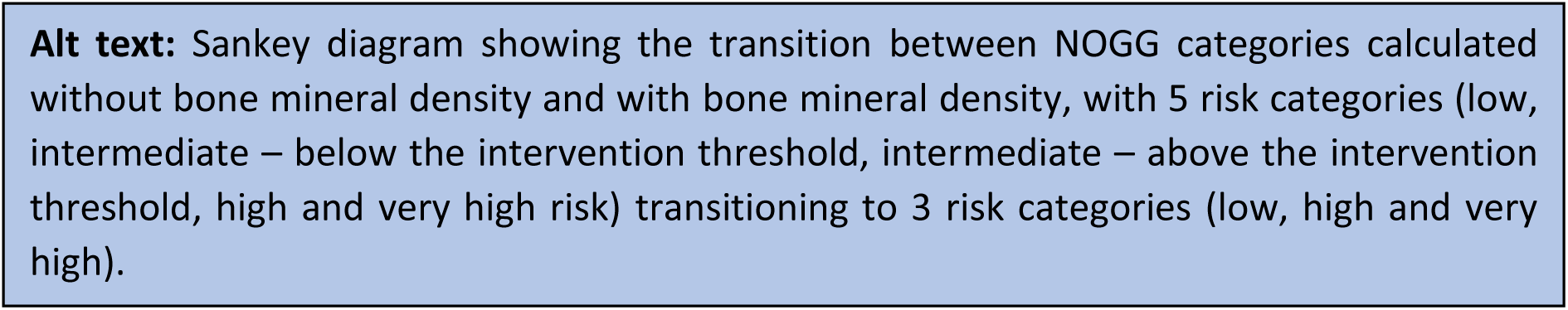

### What is the impact of recurrent falls?

Falls frequency data was collected from all 182 participants, of whom 74 (40.7%) experienced recurrent falls. Recurrent-fallers did not differ from non-recurrent-fallers in terms of age (mean difference=-0.2, p=0.887) or sex (37.8% vs 32.4% female respectively, p=0.449). However, recurrent fallers had higher MDS-UPDRS Part III (motor) scores by an average of 9.8 points (p<0.001) (Appendix.7).

After falls-adjustments (n=182) FRAX probabilities (excluding BMD) increased for MOF by mean 2.0% (p<0.001) and hip fractures by mean 1.1% (p<0.001) (Appendix.8). Thus NOGG- categories were increased for 34 people (18.7% of study population and 45.9% of recurrent fallers), leading to a change from lifestyle advice (low, or intermediate (below intervention threshold) risk) to treatment recommendation (intermediate (above intervention threshold) or high-risk) for 9 people (4.9% of study population and 12.2% of recurrent fallers), and from treatment to treatment with liaison with local bone-health specialists (very high-risk) for 9 people (4.9% of study population and 12.2% of recurrent fallers) (Figure 3.A).

**Figure 3.**
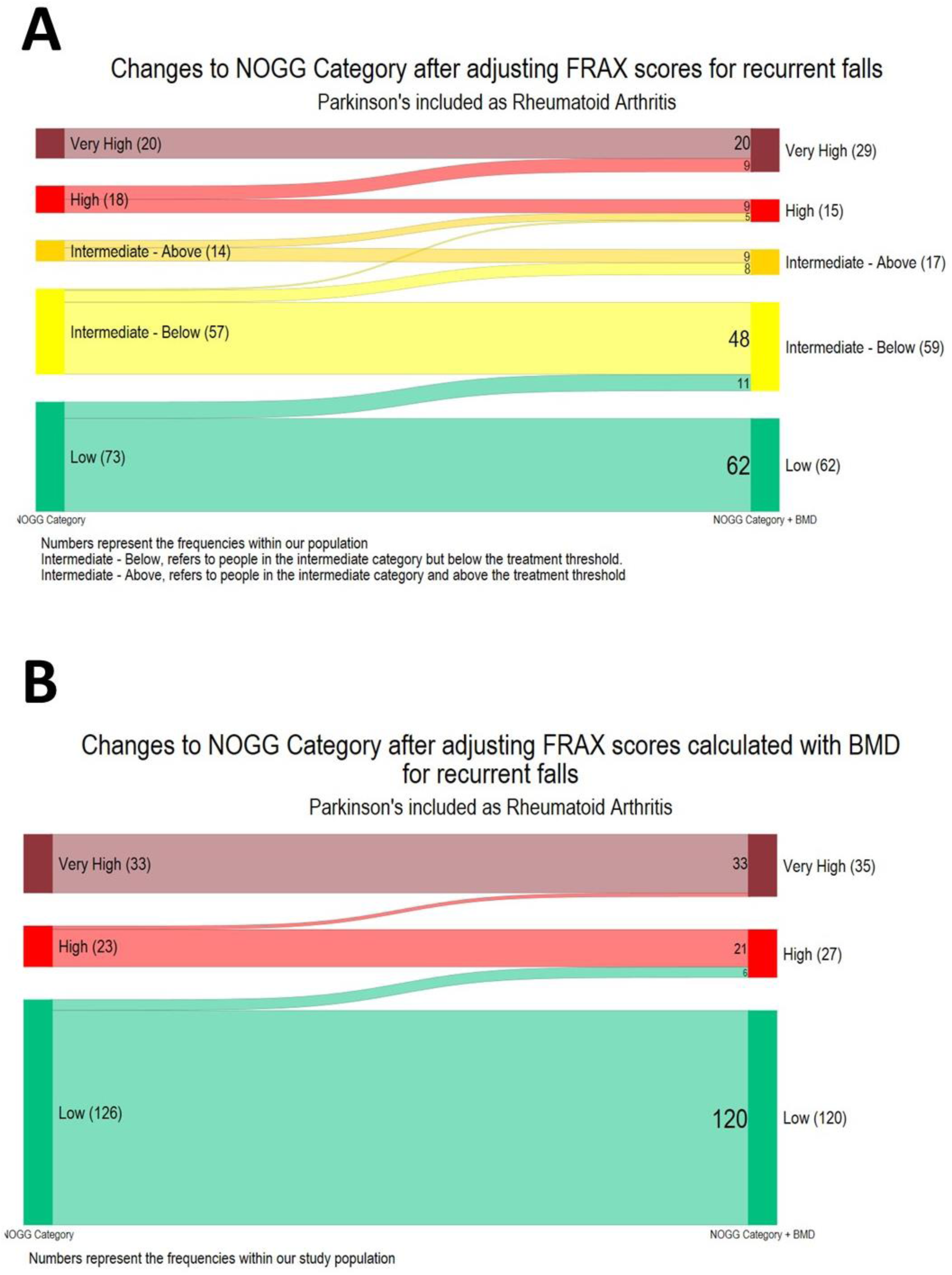
Transitions between NOGG-categories following the application of the NOGG falls adjustment A Sankey diagram to show the transition between the NOGG-categories following application of the NOGG falls adjustment to the FRAX probabilities (n=182), where PD is included as rheumatoid arthritis. The numbers on the plots represent the number of participants within each category A) FRAX without the inclusion of Bone Mineral Density, the NOGG -categories are for major osteoporotic fracture (n=182) B) FRAX with the inclusion of Bone Mineral Density, the NOGG-categories are for either hip or major osteoporotic fracture whichever was greater (n=182)

Recalculated FRAX probabilities (with BMD) were also adjusted for falls (n=182) which increased MOF probabilities by mean 1.5% (p<0.001) and hip fracture probabilities by mean 0.5%, (p<0.001) (Appendix.9). Thus NOGG-categories were increased for 8 people (4.4% of study population and 10.8% of recurrent fallers), leading to a change from lifestyle advice (low, or intermediate (below intervention threshold) risk) to treatment recommendation (intermediate (above intervention threshold) or high-risk) for 6 people (3.3% of study population and 8.1% of recurrent fallers), and from treatment to treatment with liaison with local bone-health specialists (very high-risk) for 2 people (1.1% of study population and 2.7% of recurrent fallers) (Figure 3.B).

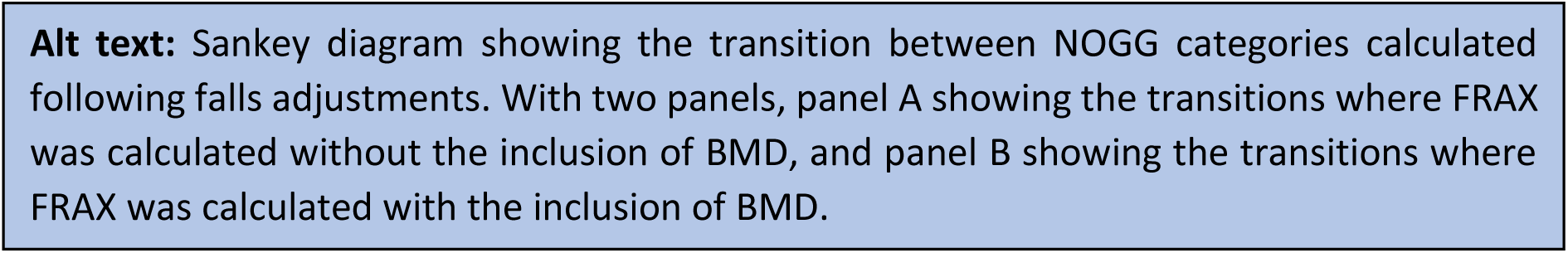

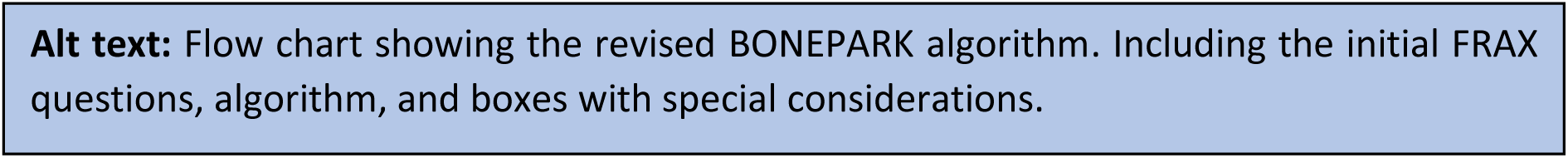

## Discussion

Despite some notable advances in the management of bone health, patients with PD continue to receive suboptimal care. To tackle this, we undertook a cross-sectional study of people with parkinsonism to determine their fracture risk using FRAX and the implications for treatment based on the NOGG recommendations. We identified the current treatment gaps and addressed these by generating an updated algorithm for the assessment and treatment of bone-health in PD (Figure 4).

**Figure 4.**
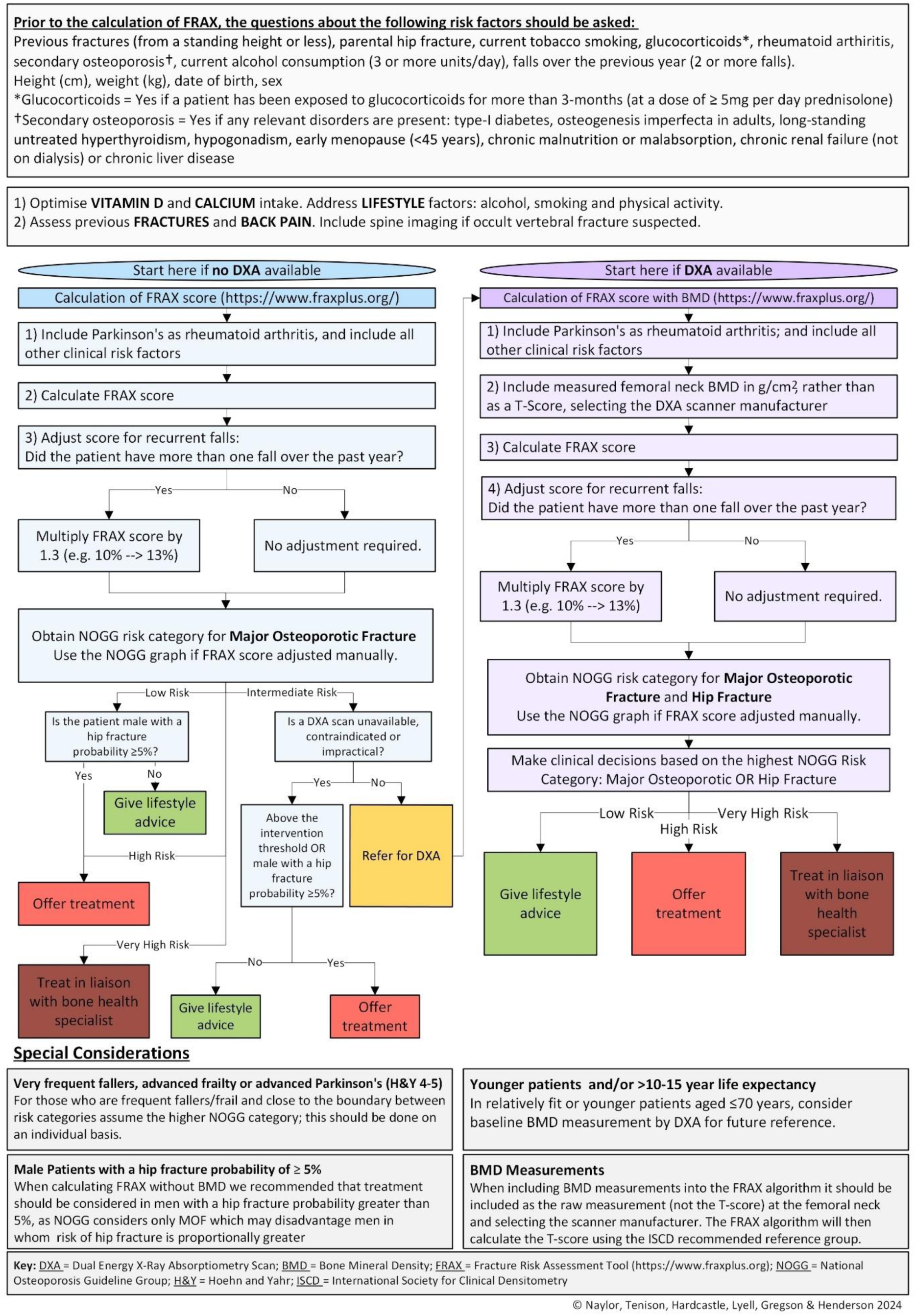
BONE PARK 2 Algorithm

Consistent with the findings of the Parkinson’s UK national audit [33], we found a substantial treatment gap within our population with only a third or less of participants receiving the NOGG-recommended treatment. Our results suggest that DXA was feasible in this population given 87% attended and we did not find that tremor, camptocormia or mobility precluded DXA scanning. Additionally, DXA was broadly reassuring as most participants (73.6%) received the same recommendation following FRAX recalculation.

Importantly, the inclusion of falls as a risk factor altered the FRAX probabilities, with an additional 5% recommended for treatment after falls adjustment.

The bone-health treatment gap is marked in PD, despite the latest NOGG guidance. This gap is also potentially greater than reported, given that fracture risk is being under-estimated within this population since PD-associated fracture risk is not fully captured in FRAX, and we explored several tenets of this. In lieu of PD being included as a risk factor in FRAX, we selected *RA* as a proxy marker [29]. Previous guidelines recommended the use of *Secondary Osteoporosis* as a proxy marker [14]; however, this approach inadvertently assumes PD-associated fracture risk is entirely mediated through low BMD, the assumption applied by FRAX to all causes of *Secondary Osteoporosis* [16]. Practically, this means that including PD as *Secondary Osteoporosis* only uplifts FRAX scores calculated without BMD. However, recent evidence suggests that PD increases fracture risk independent of BMD, which can be reflected by including PD by proxy as RA, because this uplifts FRAX score both with and without BMD [29]. However, this will still only capture some of the PD-associated fracture-risk, and ideally PD would be incorporated as a distinct risk factor into future iterations of the FRAX algorithm.

Falls are a known independent risk factor for fracture [21–23], and approximately 60% of people with PD experience falls [20]; without incorporating falls into fracture risk assessment, risk is underestimated in this group. We employed the falls adjustment from the NOGG guidance [17,31]; however, this over-simplifies the influence of falls on fracture-risk. Recent meta-analyses suggest that just one fall in the previous year confers a risk for fracture, and that hazard ratios increase with the number of falls; whilst an inflation could be applied per fall up to five falls [31], we maintained the NOGG-recommended approach [17]. Additionally, there is evidence that falls increase fracture risk to a greater extent in men, and that the increased risk from falls decreases with age [21,34]. To account for these intricate effects requires the integration of falls into FRAX; whilst a falls-adjustment is available on the FRAXplus platform [35], the associated cost precludes its use in NHS clinical practice. Until such updates to FRAX are freely available, the NOGG adjustment provides a pragmatic solution to guide treatment decisions in patients experiencing falls.

In those who are frail, very frequent fallers or close to the boundary of the higher NOGG- category, consider, on an individual basis, management according to the higher risk category. Clinical judgment is necessary regarding patient preferences towards treatment, co-morbidities and life-expectancy; and recognising that FRAX including any adjustments will not completely capture fracture risk associated with parkinsonism.

Currently two websites host FRAX risk algorithms; we recommend that the new FRAXplus website (www.fraxplus.org) be used for FRAX calculation, as the old website (https://frax.shef.ac.uk/FRAX) will soon be retired. The FRAX algorithm specifies that BMD should be measured at the femoral neck and, if BMD is included as a T-Score, this should be calculated using the NHANES III age 20-29 female reference data [27], in line with current international and national guidelines. However, in our experience, radiology departments may use alternative reference data and/or not specify the reference data used. Since the choice of reference population will affect the generated T-scores, it is advisable to enter the absolute value in g/cm^2^ for FN-BMD and select the appropriate scanner manufacturer within the FRAX tool.

Our study had both strengths and limitations. This study is strengthened by its use of a representative sample, that included participants across the Hoehn and Yahr, and clinical frailty scales, as those with more advanced disease or greater frailty are typically under- represented in research [35]. Furthermore, we were able to collect bone-health data from 99.5% of participants. There are some limitations to consider. Whilst we were able to obtain DXA scans from most participants, those who did not receive a scan (13%) were older, had worse PD symptoms and were frailer, therefore fracture risk is likely to have been underestimated compared with the wider PD population. The practicalities of DXA scanning likely precluded these individuals from taking part. As well as an additional hospital appointment, DXA scanning requires the patient to transfer to the scanning bed, lie flat, and maintain position for the scan duration. This is an important consideration, since frailty is associated with poorer outcomes following fragility fracture [37]. The latest NOGG guidance helps to mitigate this; if FRAX assessment (without BMD) produces a probability in the intermediate risk category but DXA scanning is unavailable, contraindicated or impractical, treatment can be offered to patients above the age-specific intervention threshold. We have reflected this in our proposed algorithm.

We also acknowledge that using NOGG-categories calculated without BMD to determine treatment can disadvantage men in whom risk of hip fracture is proportionally greater. Therefore, in the BONEPARK algorithm (Figure.4) we recommend that treatment also be considered in men with a FRAX hip fracture probability greater than 5% when calculating FRAX without BMD.

The ability to adjust FRAX for falls requires accurate recollection of falls over the previous year. However, both falls, and falls-related injuries are under-reported amongst older people and retrospective recall can be unreliable [38–40]. This might have resulted in an underestimation of fracture risk, although we sought to mirror how our proposed guidelines would be implemented in clinical practice.

We acknowledge that FRAX has other limitations, such as not accounting for the number or type of previous fractures. Since FRAX may underestimate fracture risk in certain individuals, such as those with multiple prior fractures, hence the algorithm reiterates the need for clinical judgement when making treatment decisions.

This revised BONE-PARK algorithm is informed by the latest NOGG guidance. We have incorporated our novel findings regarding falls adjustment and operationalised it specifically for Parkinson’s clinicians. The algorithm can be used both without and with BMD measurements and should help clinicians to assess bone health and fracture risk accurately and thenceforth make appropriate treatment decisions in tandem with their patients.

## Supporting information

All Supplementary Material

## Data Availability

The participants of this study did not give written consent for their data to be shared publicly, so due to the sensitive nature of the research supporting data is not available.

## Acknowledgements

Asjad Naqvi for the development and update of the Sankey Diagram package for Stata (https://github.com/asjadnaqvi/stata-sankey”) Jackie Shipley, and the clinical measurement team at the Royal United Hospitals NHS Foundation Trust, for performing the DXA scans. The authors would like to thank the PPI contributors who helped to inform this study, and the study participants and their carers for their participation. Mícheál Ó Breasail and Nicola Giles for organisation of the DXA scanning. Katie Lloyd & Charlotte McDonald for overall running of PRIME-RCT. Anahita Nodehi & Michael Lawton for their guidance with the statistical analysis.

## Funding

This trial was funded by Royal Osteoporosis Society (509) Parkinson’s Excellence Network (M-22-004) and Gatsby (GAT3676).

## Declarations of sources of funding

E.T. is funded by a National Institute for Health and Care Research Academic Clinical Lectureship. CLG is funded by the National Institute for Health and Care Research (NIHR302394). The views expressed are those of the authors and not necessarily those of the NIHR or the Department of Health and Social Care

## Notes

### Competing Interest Statement

The authors have declared no competing interest.

### Clinical Trial

NCT05127057

### Author Declarations

Ethical approval was granted by London-Harrow Research Ethics Committee (REC reference 21/LO/0387) on 14/7/2021.

