## Supplementary material for "Assessing and managing bone-health and fracture risk in Parkinson’s disease: the BONE PARK 2 protocol": All Supplementary Material

**Appendix 1** A flow chart to demonstrate how T-score site was chosen, and how osteoporosis, osteopenia (low bone mass) and normal BMD were categorised.

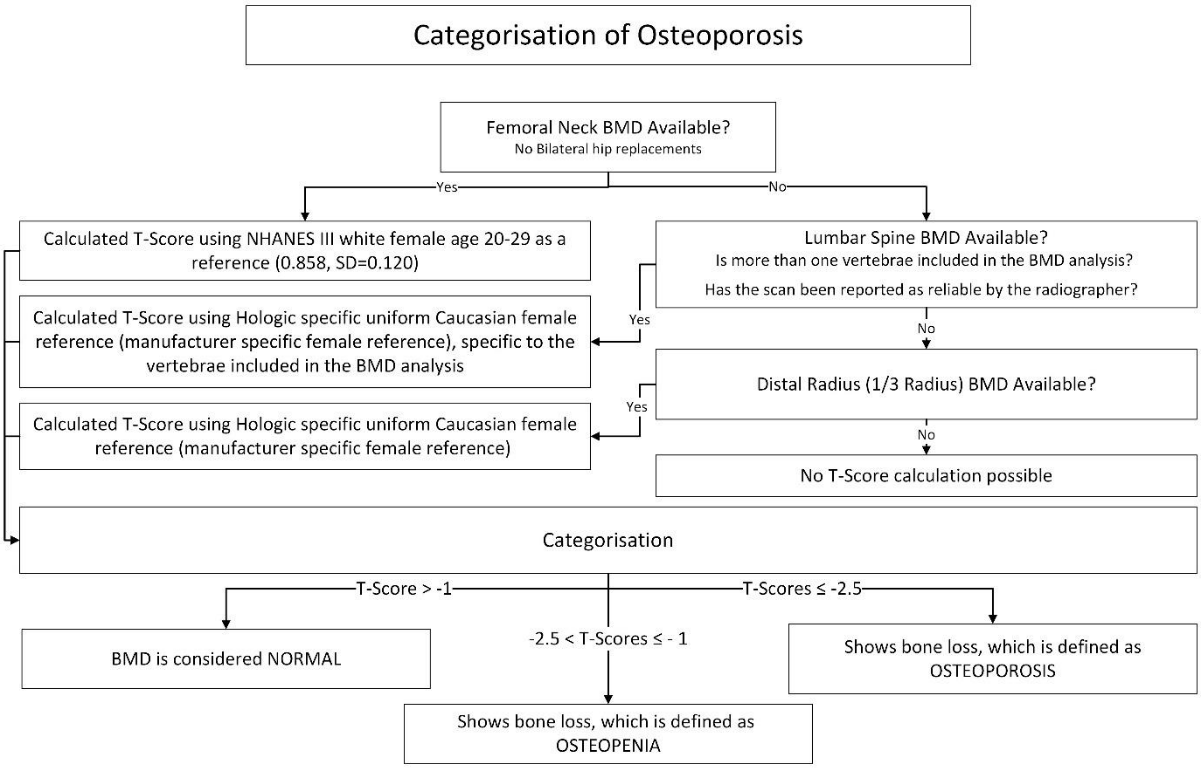

**Appendix 2** How zoledronate and denosumab were considered in the medication gap analysis.

Participants were considered ‘on treatment’ if anabolic or anti-resorptive treatments were started and not discontinued before the baseline visit. Treatment with Zoledronate was considered to be discontinued if participants had their most recent intravenous infusion more than 18 months before the baseline visit. Denosumab treatment was considered to be discontinued if participants had their most recent injection more than 8 months before the baseline visit.

### **Appendix 3** Reasons given by participants for not having a DXA scan

| **Reason** | N | Percentage |
|---|---|---|
| Back pain | 1 | 4% |
| Burden | 1 | 4% |
| DNA/Cancelled | 5 | 19% |
| Housebound or bedbound | 3 | 11% |
| Mobility | 2 | 7% |
| No specific reason given | 9 | 33% |
| Additional travel | 4 | 15% |
| Uncertainty about scan benefits | 1 | 4% |
| Withdrawn from PRIME-RCT | 1 | 4% |
| **Total** | 27 | 100% |

### **Appendix 4** Comparing *age, sex, MDS-UPDRS Part III and CFS Scores of* those who did and did not have a DXA scan

| | No DXA | DXA Scan Available | p-value |
|---|---|---|---|
| N | 28 (13.1%) | 186 (86.9%) | |
| Age at Visit 1 | 78.6 (5.8) | 73.9 (8.3) | 0.005 |
| Gender | | | |
| Female | 11 (39.3%) | 64 (34.4%) | 0.614 |
| Male | 17 (60.7%) | 122 (65.6%) | |
| MDS-UPDRS Section 3 | 58.6 (21.9) | 40.3 (18.5) | <0.001 |
| Frailty | | | |
| CFS < 5 | 5 (17.9%) | 127 (68.3%) | <0.001 |
| CFS ≥ 5 | 23 (82.1%) | 59 (31.7%) | |

Continuous data are described as mean (SD), and compared with t-test
Categorical data is described as frequency (Percent%), and compared with Chi Square test
Frailty is described as frequency (Percent%), and compared with a z-test of proportions

**Appendix 5** Frequency of prescribed Bone Protective Medications in our study population

| | Frequency |
|---|---|
| Total N | 182 |
| None | 161 (88.5%) |
| Alendronate | 16 (8.8%) |
| Risedronate | 3 (1.6%) |
| Zoledronate | 1 (0.5%) |
| Denosumab | 1 (0.5%) |

**Appendix 6** FRAX-derived probabilities of fracture, and NOGG risk categorisation, before and after the inclusion of Femoral Neck BMD in the FRAX algorithm

| | Femoral Neck BMD Included | |
|---|---|---|
| | No | Yes |
| N | 182.0 (50.0%) | 182.0 (50.0%) |
| FRAX probability of MOF* | 16.3 (11.1) | 12.7 (9.0) |
| FRAX probability of Hip fracture* | 8.5 (9.0) | 5.0 (6.8) |
| NOGG Category* | | |
| Low Risk | 73.0 (40.1%) | 126.0 (69.2%) |
| Intermediated Risk - below IT† | 57.0 (31.3%) | 0.0 (0.0%) |
| Intermediate Risk-above IT‡ | 14.0 (7.7%) | 0.0 (0.0%) |
| High Risk | 18.0 (9.9%) | 23.0 (12.6%) |
| Very High Risk | 20.0 (11.0%) | 33.0 (18.1%) |

Continuous data are described as mean (Standard Deviation)
Categorical data is described as frequency (Percent%)
* PD was included as rheumatoid arthritis
† Intermediate Risk, but below the intervention threshold
‡ Intermediate Risk, but above the intervention threshold

### **Appendix 7** Comparing those who did and did not experience recurrent falls

| | No Recurrent Falls | Recurrent Falls | p-value |
|---|---|---|---|
| N | 108 (59.3%) | 74 (40.7%) | |
| Age | 73.8 (8.1) | 73.9 (8.5) | 0.887 |
| Sex | | | |
| Female | 35 (32.4%) | 28 (37.8%) | 0.449 |
| Male | 73 (67.6%) | 46 (62.2%) | |
| MDS-UPDRS Part III | 36.2 (18.3) | 46.1 (17.1) | <0.001 |

Continuous data are described as mean (SD), and compared using a t-test
Categorical data is described as frequency (Percent%), and compared using a Chi square test

### **Appendix 8** FRAX-derived probabilities of fracture without the inclusion of femoral neck BMD, before and after adjustment for recurrent falls

| | No Falls Adjustment | Falls Adjustment Applied* |
|---|---|---|
| N | 182 (50.0%) | 182 (50.0%) |
| FRAX Score for MOF | 16.3 (11.1) | 18.3 (12.8) |
| FRAX Score for Hip | 8.5 (9.0) | 9.6 (10.1) |

Continuous data are described as mean (SD)
*Falls adjustment is a 30% increase in FRAX probability applied where people experienced recurrent falls (2 or more falls over the previous year) – there were 74 recurrent fallers in the study population

### **Appendix 9** FRAX-derived probabilities of fracture with the inclusion of femoral neck BMD, before and after adjustment for recurrent falls

| | No Falls Adjustment | Falls Adjustment Applied* |
|---|---|---|
| N | 182.0 (50.0%) | 182.0 (50.0%) |
| FRAX Score for MOF | 12.7 (9.0) | 14.2 (9.9) |
| FRAX Score for Hip | 5.0 (6.8) | 5.5 (7.2) |

Continuous data are described as mean (SD)
*Falls adjustment is a 30% increase in FRAX probability applied where people experienced recurrent falls (2 or more falls over the previous year) – there were 74 recurrent fallers in the study population
